# Genome-wide contribution of common Short-Tandem Repeats to Parkinson’s Disease genetic risk

**DOI:** 10.1101/2021.07.01.21259645

**Authors:** Bernabe I. Bustos, Kimberley Billingsley, Cornelis Blauwendraat, J. Raphael Gibbs, Ziv Gan-Or, Dimitri Krainc, Andrew B. Singleton, Steven J. Lubbe, For the International Parkinson’s Disease Genomics Consortium (IPDGC)

## Abstract

Parkinson’s disease (PD) is a complex neurodegenerative disorder with a strong genetic component, where most known disease-associated variants are single nucleotide polymorphisms (SNPs) and small insertions and deletions (Indels). DNA repetitive elements account for >50% of the human genome, however little is known of their contribution to PD etiology. While select short tandem repeats (STRs) within candidate genes have been studied in PD, their genome-wide contribution remains unknown. Here we present the first genome-wide association study (GWAS) of STRs in PD. Through a meta-analysis of 16 imputed GWAS cohorts from the International Parkinson’s Disease Genomic Consortium (IPDGC), totalling 39,087 individuals (16,642 PD cases and 22,445 controls of European ancestry) we identified 34 genome-wide significant STR loci (p < 5.34×10^-6^), with the strongest signal located in *KANSL1* (chr17:44205351:[T]_11_, p=3×10^-39^, OR=1.31 [CI 95%=1.26-1.36]). Conditional-joint analyses suggested that 4 significant STRs mapping nearby *NDUFAF2, TRIML2, MIRNA-129-1* and *NCOR1* were independent from known PD risk SNPs. Including STRs in heritability estimates increased the variance explained by SNPs alone. Gene expression analysis of STRs (eSTR) in RNASeq data from 13 brain regions, identified significant associations of STRs influencing the expression of multiple genes, including PD known genes. Further functional annotation of candidate STRs revealed that significant eSTRs within *NUDFAF2* and *ZSWIM7* overlap with regulatory features and are associated with change in the expression levels of nearby genes. Here we show that STRs at known and novel candidate PD loci contribute to PD risk, and have functional effects in disease-relevant tissues and pathways, supporting previously reported disease-associated genes and giving further evidence for their functional prioritization. These data represent a valuable resource for researchers currently dissecting PD risk loci.

## INTRODUCTION

Parkinson’s disease (PD) is a complex neurodegenerative disease with an established genetic component. Studies over the years have identified several rare variants that cause or significantly increase the risk of disease in carriers, and genome-wide association studies (GWAS) have recently uncovered 90 common variants that influence PD risk^1^. It is estimated that common GWAS variants account for 16-36% of the overall genetic heritability of PD^1,2^ highlighting that a large proportion of the missing heritability remains to be identified.

The vast majority of PD genetics studies have focused on the role of single nucleotide polymorphisms (SNPs), meaning that contributions of other genetic elements such as structural variants and repetitive elements have largely been ignored. Repetitive elements represent more than 55% of the human genome^3^. Short tandem repeat expansions (STRs), are small repetitive units ranging from 1 to 7 base pairs in length that vary among individuals, and account for ∼10% of all repetitive elements^4^. STRs are the cause of several neurological diseases and are associated to genes such as in Fragile X syndrome (*FMR-1*)^5,6^, Huntington’s disease (*HTT*)^7^, amyotrophic lateral sclerosis and frontotemporal dementia (*C9ORF72)*^*8,9*^, and spinocerebellar ataxia (*SCA1*)^10^, and have also been linked to numerous complex neurological and psychiatric traits^11^. A role for STRs as drivers of GWAS signals have been identified^12^, where a risk SNP connected adjacent GGAA repeats by converting an interspaced GGAT motif into a GGAA motif, thereby increasing the number of consecutive GGAA motifs and modifying the activity of its sequence and functional impact. STRs have also been shown to significantly regulate gene expression and contribute to phenotypic plasticity^13^. STRs therefore represent a potential source of unexplored genetic variation that may account for some of the missing heritability of PD. In this regard, other repetitive elements, such as satellite repeats, have been shown to alter gene expression in blood of PD patients^14^. However, no genome-wide assessment of STRs in large population studies has yet been performed in this disease. Due to their more complex and highly repetitive structure compared to SNVs, STRs have been difficult to assess. Despite the recent explosion of genetic data stemming from next generation sequencing, STRs are still difficult to genotype. Recent advances in PCR-free deep sequencing methods and STR genotyping tools now allow for the simultaneous assessment of STRs genome-wide^15^. Studies have shown high linkage disequilibrium (LD) between STRs and SNPs across the genome^16^. Exploiting this high LD, Saini *et al*. (2018)^17^ generated a phased SNP-STR haplotype panel based on the 1000 Genomes Project samples that allows for the accurate genome-wide imputation of common STRs into array-based genotype data. To assess the role of common STRs in PD risk, we imputed and interrogated STRs across 16 independent PD case-control cohorts, totaling 39,087 individuals available through the International Parkinson’s disease Genomics Consortium (IPDGC).

## RESULTS

### Meta-analysis of IPDGC GWAS cohorts imputed with an STR reference panel

The 16 GWAS cohorts used in this study, with a combined sample size of 39,087 individuals composed of 16,642 PD cases and 22,445 controls of self-reported European ancestry (**Supplementary Table 1**). After cohort-wise quality controls (see Methods), we performed genome-wide imputation using the 1000 Genomes STR-SNP reference panel^17^, and carried out case-control association analyses with PD status following a meta-analysis of fixed effects across all cohorts. After removing variants with high heterogeneity across meta-analyses (I^2^ >0.8), we obtained association p-values for 407,879 STRs, where 214 variants surpassed the threshold for genome-wide significance of 5.34×10^-6^ (**Figure 1, upper side)**, which was estimated by permutation procedures for the STR reference panel, as described elsewhere^18^. The inflation factor lambda for the association was 1.18 and the rescaled lambda for 1000 cases and controls (λ 1000) was 1.01. To characterize and identify independently associated STRs, we first performed a conditional-joint analysis using GCTA-COJO^19^ and identified 34 STR variants mapping to 32 unique nearby genes, with the strongest signal located in *KANSL1* (chr17:44205351:[T]_11_, p=3×10^-39^, OR=1.31 [CI 95%=1.26-1.36]), and followed by *SNCA* (chr4:90662073:TATTT[GT]_8_AT[GT]_7_, p=3.36×10^-25^, OR=1.36 [CI 95%=1.28-1.45]) (**Table 1**). Since STRs were imputed by leveraging LD information from SNPs, we carried out a secondary GCTA-COJO analysis including the meta-analys results from imputed, filtered SNPs (**Figure 1, lower side**), obtaining a total number of 8,179,378 SNPs and STRs in all cohorts. We found eight loci with associations led by STRs (**Supplementary Table 2**), and in order to refine these results, we further investigated their LD patterns with the 90 known PD risk variants from the 2019 PD GWAS meta-analysis^1^, and found that four of the eight STRs had LD r^2^ <0.5 with any of the known PD variants (**Supplementary Table 3**), indicating that these could be potential new PD risk signals:

- a tetranucleotide repeat within the 3^rd^ intron of *NDUFAF2* (risk allele chr5:60437492:AA[TGAA]_7_, p=6.49×10^-8^, OR=1.30, CI 95%=1.18-1.43) (**Figure 2A**);
- a mononucleotide repeat downstream of *TRIML2* (risk allele chr4:189000404:TT[A]_12_, p=1.44×10^-7^, OR=1.31, CI 95%=1.19-1.44) (**Figure 2B**);
- a mononucleotide repeat downstream of *MIR129-1* (risk allele chr7:127793488:[T]_15_G, p=2.79×10^-7^, OR=1.16, CI 95%=1.09-1.23) (**Figure 2C**); and
- a mononucleotide repeat within the 44^th^ intron of *NCOR1* (risk allele chr17:15941750:[T]_11_, p=3.77×10^-6^, OR=1.08, CI 95%=1.04-1.12) (**Figure 2D**).

**Table 1.**
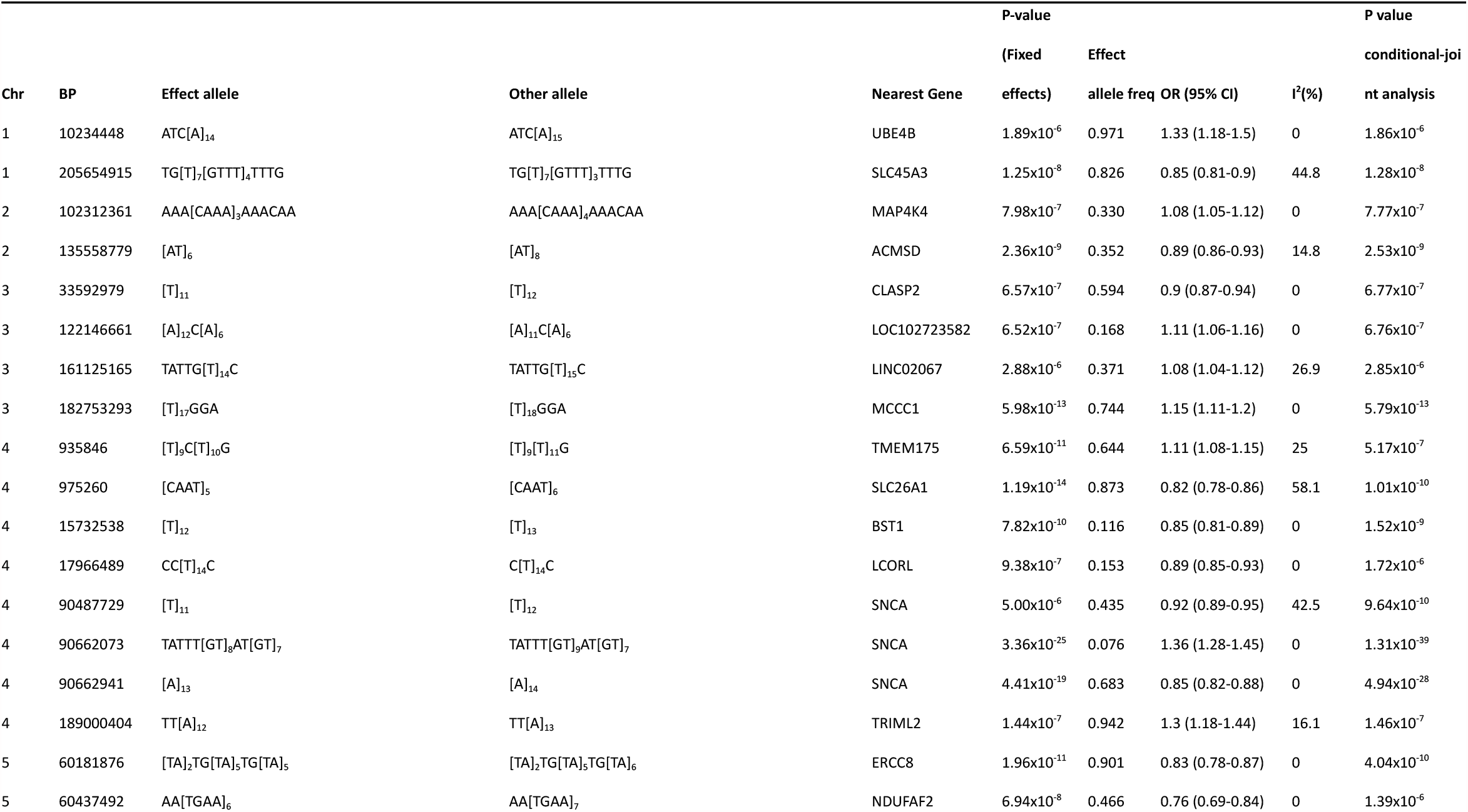
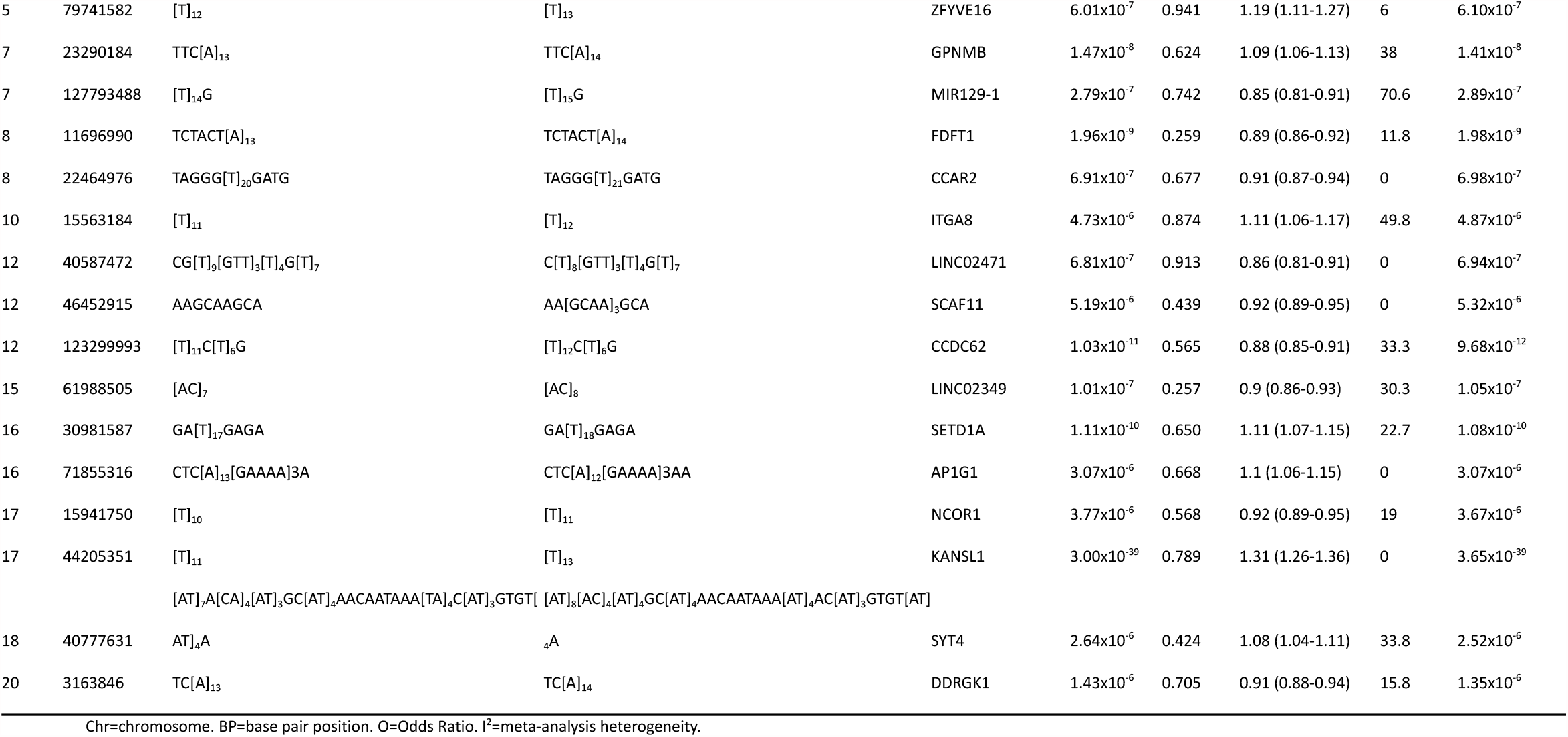
**Genome-wide significant STR loci from meta-analysis of 16 case-control Parkinson’s Disease GWAS cohorts**.

**Figure 1.**
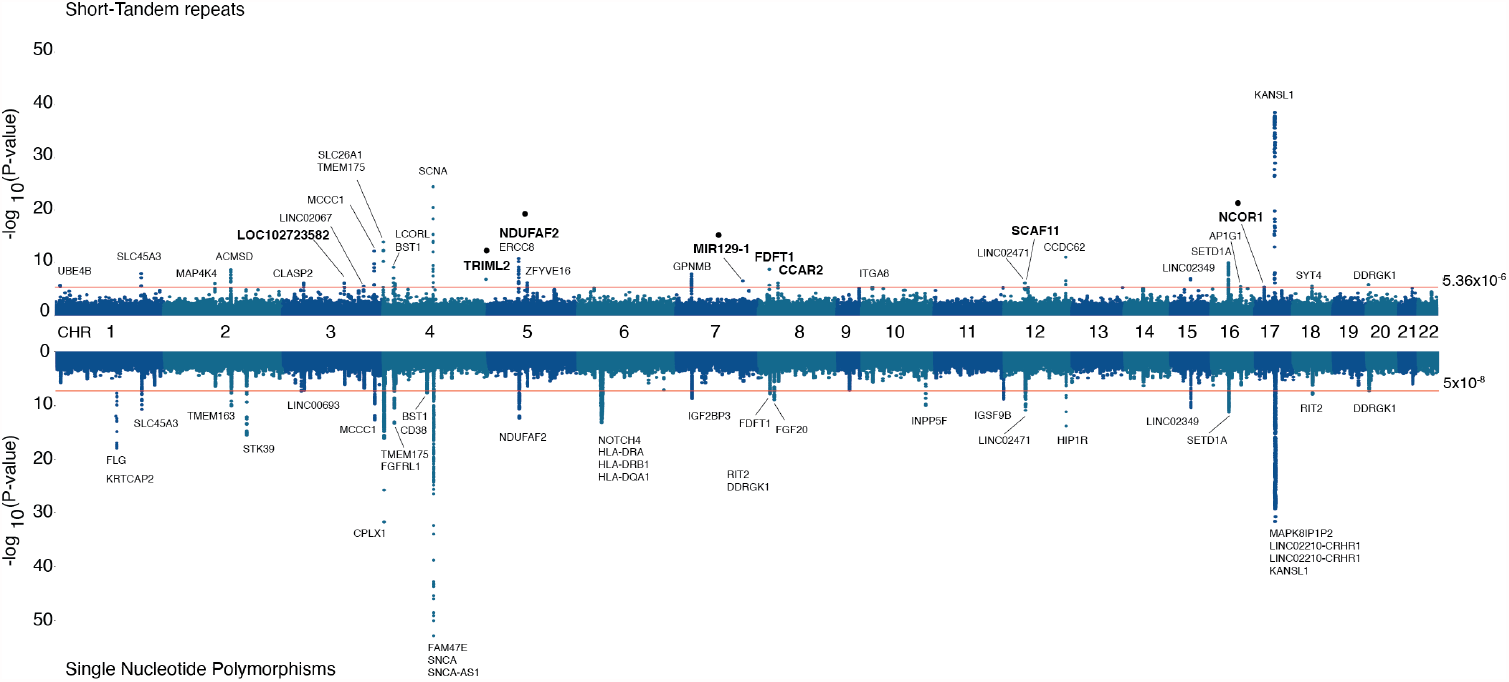
Genome-wide association results for imputed STRs and SNPs in 16 PD GWAS cohorts from the International Parkinson’s disease genomics consortium. Hudson plot representing the association analysis results for STRs (upper) and SNPs (lower) across the human genome, showing the 34 genome-wide significant STR loci (p<5.34×10^-6^) after the conditional-joint analysis with GCTA. Genes in bold represent the loci influenced by STRs after including SNPs associations. Genes with a black dot at the top represent STR loci independent from the current 90 PD risk variants from the 2019 PD GWAS meta-analysis.

**Figure 2.**
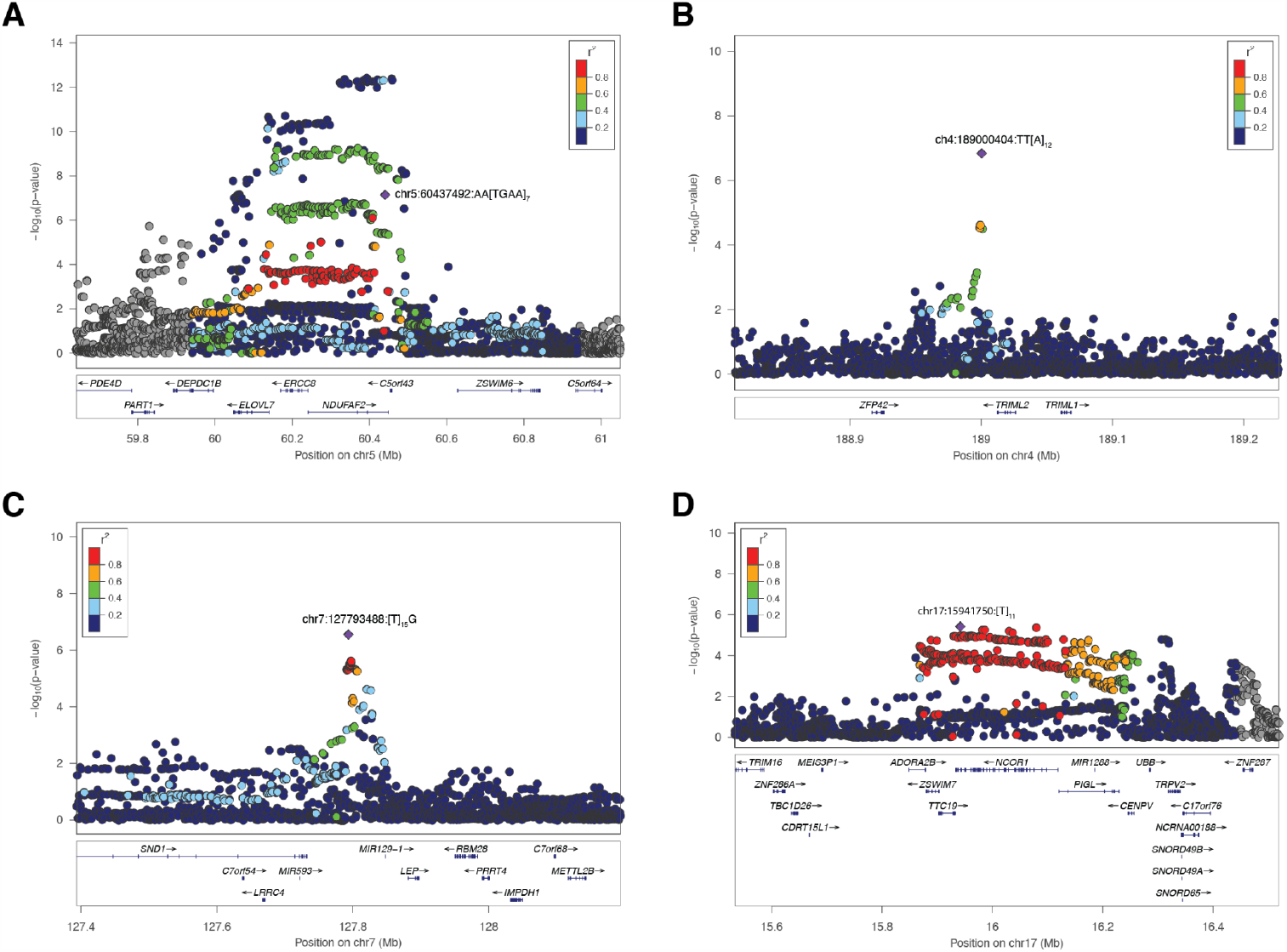
Regional association plots for the 4 candidate independent STR loci. Locus zoom plots were generated for the 4 GCTA-nominated independent STR loci from SNPs and 90 risk PD loci in **(A)** chromosome 5 within *NDUFAF2*, **(B)** chromosome 4 nearby *TRIML2*, **(C)** chromosome 7 nearby *MIR129-1* and **(D)** chromosome 17 within *NCOR1*. Lead STR variant is depicted as a purple diamond and nearby variants (STRs and SNPs) in circles colored by their LD r^2^ value to the lead STR variant. Gene annotations for each region are displayed in the bottom part of each panel, showing gene strand orientation with arrows.

It is important to note here that the independent STR signal at *NDUFAF2* (chr5:60437492:AA[TGAA]_7_) is within a known PD risk locus (mapping to *ELOVL7*), and was previously identified through Mendelian randomization to be significantly associated with risk of PD^1^. Moreover, further LD analysis on this locus showed a high D’ statistic with the closest known PD risk SNP at that locus (D’=0.94 with rs1867598) indicating that, regardless of frequency disparities, the independency suggested by the GCTA-COJO analysis should be taken with caution.

### Quantifying the heritability of STRs in PD

The genetic heritability of PD was recently estimated to be 22%^1^. Here, assuming a global disease prevalence of 0.2%^2^, we leveraged the GCTA-LDMS method^19^ and estimated that common STRs (MAF >1%) account for 15.2% (SE=0.01) of the additive heritability of the disease on the liability scale. Heritability for imputed SNPs in the same data accounted for 26.9% (SE=0.02), similarly to what was obtained in Keller *et al*., 2012 using GCTA as well. After including both common STRs and SNPs in the analysis, the heritability estimate increased to 28.8% (SE=0.02). This increase of 1.9% in the heritability estimate due to common STRs corresponds to a 7% increase from the SNP based estimate.

### eSTR analysis

We functionally assessed the impact of the 34 significant STR associations through an expression quantitative trait loci analysis (eSTR). We investigated each locus extracting the leading STR and other STRs in high LD (r^2^ >0.5) within 1 Mb up- and downstream, obtaining 105 variants for further analysis (**Supplementary Table 4**). We used normalized gene expression data from frontal cortex from the North American Brain Expression Consortium (NABEC)^20^, and 13 brain tissues from the Genotype-Tissue Expression Consortium (GTEx v.8)^21^, and identified 10,252 STR-gene associations for both datasets and all tissues (**Figure 3A**). Of these, 840 associations showed a False discovery rate (FDR) corrected p<0.05, corresponding to 234 unique eGenes (genes with at least one significant variant), that included 19 of the 78 loci identified in the 2019 PD GWAS meta-analysis (genes nominated from the 90 PD risk variants): *RIT2, TMEM163, MCCC1, LCORL, CTSB, SETD1A, CRHR1, GPNMB, BIN3, TMEM175, GAK, MAP4K4, SNCA, SPTSSB, WNT3, KPNA1, ITGA8, BST1* and *HIP1R* (**Supplementary Table 5**).

**Figure 3.**
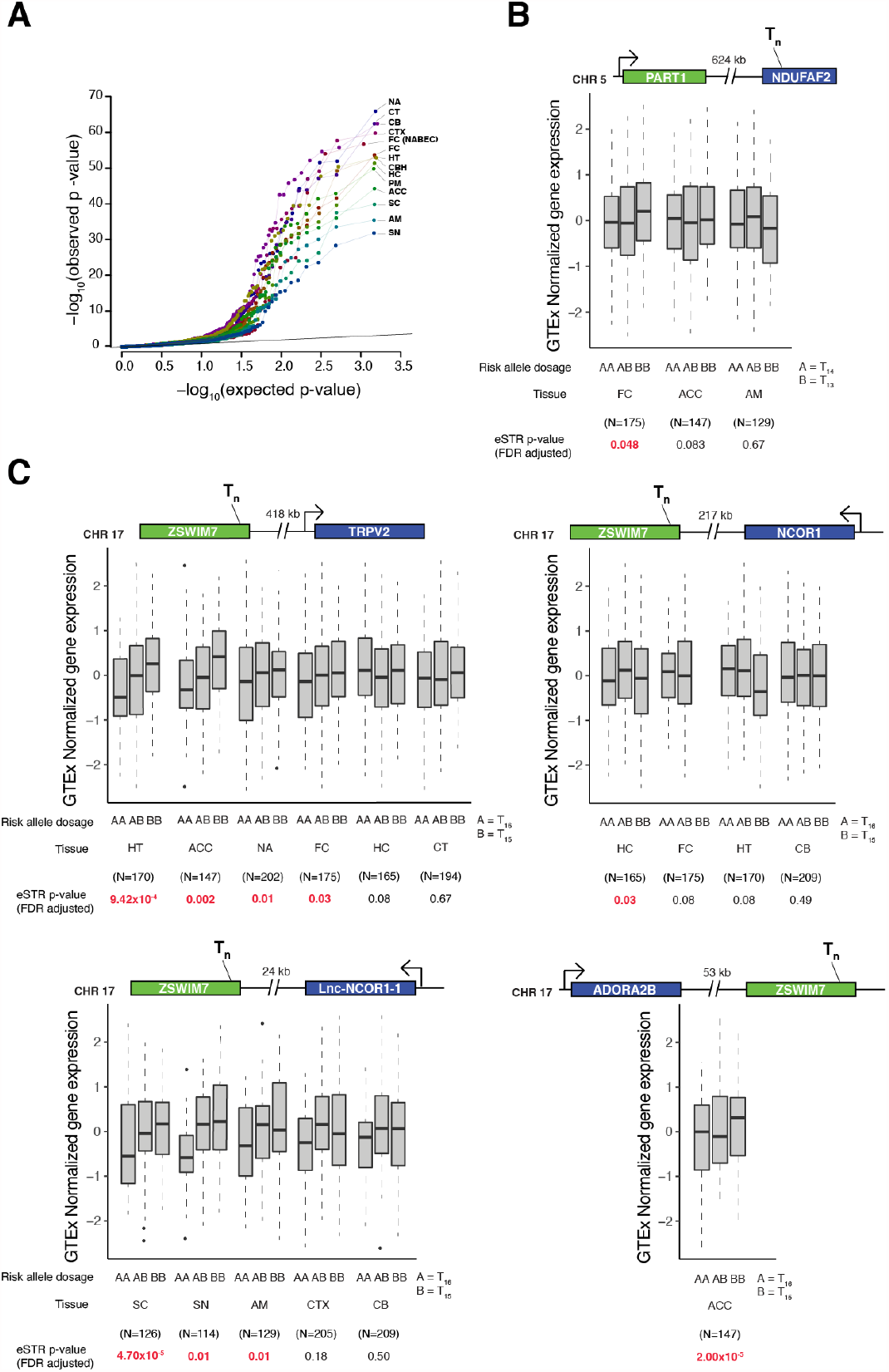
eSTR analysis of top STR loci in gene expression data from brain tissues. **(A)** Quantile-quantile plot for eSTR analysis of all 34 top loci from meta-analysis and high LD STRs, totalling 105 variants, across brain tissues from the NABEC and GTEx datasets. Colors for each dot and lines were added to enhance resolution. **(B)** Box plots showing gene expression changes associated with STR variant chr5:60408714:TC[T]_14_GTATC, across the tissues where the variant was analyzed. **(C)** Box plots showing gene expression changes associated with STR variant chr17:15902070:[T]_16_GA. Locus representation is shown at the top of each box plot, representing the STR location and the distance (in kilobases) and orientation of the target gene. At the bottom, allele dosages, tissue, sample size and eSTR FDR-adjusted p-value is shown, with p-values in red representing significant associations. NA=Nucleus accumbens (basal ganglia); CT=Caudate (basal ganglia); CB=Cerebellum; CTX=Cortex; FC (NABEC)=Frontal cortex; FC=Frontal cortex (BA9); HT=Hypothalamus;CBH=Cerebellar hemisphere; HC=Hippocampus; PM=Putamen (basal ganglia); ACC=Anterior cingulate cortex (BA24); SC=Spinal cord (cervical c-1); AM= Amygdala; SN=Substantia nigra.

To obtain functional and gene expression insights on the four candidate STRs signals obtained from our STR meta-analysis, we similarly extracted the four variants and their surrounding high LD STRs, obtaining 13 unique variants. We functionally annotated them, using regulatory features for gene expression from the Encyclopedia of DNA Elements (ENCODE)^22^, and found two STRs overlapping enhancers, transcription factor binding sites and histone marks for active transcription (*H3K4me3, H3K9Ac, H3K4me1, H3K27Ac, H3K36me3* and *H3K79me2*): one nearby *NDUFAF2* (chr5:60408714:TC[T]_14_GTATC) in high LD with the leading GWAS STR at that locus (chr5:60437492:AA[TGAA]_7,_ r^2^=0.95); and one within *ZSWIM7* (chr17:15902070:[T]_16_GA), similarly, in high LD with the leading GWAS STR for that locus (chr17:15941750:[T]_11,_ r^2^=0.84) (**Supplementary Table 6**). The PD risk allele for the eSTR in *NDUFAF2* (major allele with 13 T repetitions) is significantly associated with higher expression levels of the gene *PART1* (∼624 kb upstream) in the frontal cortex (**Figure 3B**). Interestingly, the significant eSTR in *ZSWIM7* showed associations in more brain tissues, where the risk allele in the STR meta-analysis (minor allele with 15 T repeats) was correlated with lower expression levels of *TRPV2* in the hypothalamus, anterior cingulate cortex, nucleus accumbens and frontal cortex; higher expression levels of *NCOR1* in the hippocampus; higher expression levels of *ADORA2B* in the anterior cingulate cortex; and lower expression levels of a long non-coding RNA gene located nearby *NCOR1* (CTC-529I10.1 or lnc-NCOR1-1) in the spinal cord and substantia nigra (**Figure 3C**).

### Gene-wise, gene-set and pathway enrichment analysis of PD associated STRs

MAGMA^23^ gene-wise enrichment analysis of the STR meta-analysis results yielded 47 genes surpassing genome-wide significance (Bonferroni p<2.99×10^-6^, α=0.05/16,696); **Supplementary Table 7**). Of the 47 genes, 12 overlapped with the STR meta-analysis results, 8 overlapped with 78 PD loci nominated from the 90 PD risk variants (2019 PD GWAS meta-analysis), and 27 genes have not previously been identified as enriched genes. Gene-property analysis using gene expression data from GTEx v.8, as described in FUMA^24^, showed significant enrichment of genes in the pituitary and brain tissues after FDR correction (FDR p<0.05) for the 30 GTEx general tissues, (**Supplementary Figure 2A**) and in the cerebellum, cortex, pituitary, cerebellar hemisphere and frontal cortex for the 54 GTExp specific tissues (**Supplementary Figure 2B**). We further investigated gene connectivity via protein-protein interactions using a list of 445 genes surpassing nominal gene-wise STR enrichment (MAGMA p<0.01) with WebGestalt^25^, and leveraged the Network-Topology Analysis (NTA), finding 16 subnetworks (**Supplementary Figure 3**), which were significantly enriched in 27 gene ontology categories, such as synaptic vesicle cycle (GO:0099504), presynaptic endocytosis (GO:0140238) and autophagy (GO:0006914) (**Supplementary Table 8**).

## DISCUSSION

In the present study we performed a genome-wide meta-analysis of STRs in 16 cohorts from the IPDGC. We have shown that associated STR signals overlap with known PD risk loci, and with candidate novel signals, that represented by STRs independent from current 90 risk variants^1^, and are located nearby *TRIML2, NDUFAF2, MIR129-1* and *NCOR1* (on chromosomes 4, 5, 7 and 17 respectively). We also assessed the functional consequences of the STRs at a gene expression level in brain tissues, which further supports their candidacy for functional studies to further understand the biological mechanisms behind their associations.

The fact that 88% (30/34) of the associated STRs overlap with the current list of PD GWAS risk variants is not surprising as the STRs were imputed based on their existent LD with SNPs. Known PD loci with STR associations could potentially help to explain the current unknown molecular mechanisms underlying those regions, such as in *MAPT* and *SNCA*, where evidence has shown that repetitive elements play a major role in gene expression regulation, splicing, and hence protein structure^26,27^. This overlap is further reflected in the heritability estimates we obtained which indicated that the contribution of STRs to the genetic variance of PD is largely explained by their high LD with SNPs. However, STRs have shown to increase the contribution to overall SNP-only heritability estimates especifically on gene expression^13,28^, where STRs explained between 10%–15% of the *cis*-heritability, thereby supporting our observation that STRs contribute to the heratibility of PD.

The eSTR colocalization analyses, using available RNAseq datasets, where we analyzed the top 34 STR signals and their surrounding high LD STRs, showed us different distributions of STR associations throughout the various brain regions, and at the gene level, we observed significant associations in 19 known PD risk genes, suggesting that these STRs are likely to be functionally relevant in these loci. Further investigation of the four independent nominated STRs managed to uncover likely functional mechanisms underlying the STR association in genes nearby *NDUFAF2* and *NCOR1*, due to the STR colocalization with regulatory features (epigenetic marks) involved in active transcription. The eSTR near *NDUFAF2* was found to significantly increase the expression *PART1* (**Figure 3B**). *PART1* is a long non-coding RNA that was found to be differentially expressed (downregulated) in a microarray-based analysis of 50 PD patients compared to 22 healthy controls^29^. The *ZSWIM7* eSTR was associated with significant effects on gene expression in different genes, such as *TRPV2*, a cation channel part of the Transient receptor potential family of proteins (TRPs) that are activated by physical and chemical stimuli^30^, and that are known to be involved in the regulation of ionic homeostasis, which is disrupted in PD^31^; *ADORA2B* is an adenosine receptor which has been associated with neurodegenerative conditions such as Huntington’s disease^32^, however no link to PD has been established so far; *lnc-NCOR1-1* and *NCOR1* (Nuclear Receptor Corepressor 1) are located within the same chromosomal region (short arm of chromosome 17) and were also influenced by the eSTRs. The former long non-coding gene has not been thoroughly characterized, therefore little is known about its function. The latter encodes a transcriptional inhibitor that has been found to regulate mitochondrial function^33^. Moreover, gene expression analyses showed that *NCOR1* is significantly upregulated in the substantia nigra of PD patients^34^.

This evidence suggests that those genes associated with eSTRs in PD would be good candidates for follow-up analyses.

The functional consequences of STRs captured by gene-wise and pathway analyses demonstrated that STRs are enriched in known PD-relevant pathways such as synaptic vesicle trafficking^35^ and autophagy^36^, and in tissues, such as the cortex, cerebellar hemisphere and frontal cortex. Also highlighted is the pituitary gland, that is known to express the dopaminergic receptors D2 and D4^37^ and is part of the hypothalamic–pituitary–thyroid axis, where alterations in its balance has been shown to increase risk to PD^38^.

This study marks the first (to our knowledge) PD STR GWAS to date and highlights the importance of incorporating other forms of genetic variation, such as STRs, into routine genetic analyses. Despite this, like with any study profiling repeat-based variants using short-read sequencing data, the analyses presented in this study have several limitations. First, focusing on the STR calls, STRs were imputed using a reference panel that was generated by the STR caller hipSTR using short-read whole-genome sequencing (WGS)^17^. There are two main drawbacks to this approach: (1) hipSTR cannot call STRs that are longer than the read length. Given that many of the known pathogenic STRs in neurological diseases are large repeat expansions, we currently lack the power to detect this important and potentially disease-associated class of tandem repeats; (2) As highlighted in the original study, imputation accuracy varies widely across STR loci, with highly variable multi-allelic STR only achieving ∼70% concordance. Hence future studies that validate the PD associated STRs with methods such as long-read sequencing will be crucial to confirm these loci and will be key to resolving complex repeat-based PD associated haplotypes. Second, although the majority of the STRs tested were biallelic, multi-allelic variants were split into biallelic for the GWAS and downstream analyses. This approach enabled us to perform commonly used GWAS methods for the different analysis presented in the study, but set aside the consideration of variant length as unit of analysis, an important aspect of repetitive elements, that need to be addressed in future developments with association tools that can incorporate these multi-allelic variants, which will likely give valuable insight into the specific role of repeat copy number to risk of disease we. Finally, it is important to highlight that, despite the fact that the STR panel used to impute our PD GWAS cohorts showed high levels of concordance (96.7%)^17^ with read-based callers such as hipSTR and TREDPARSE, the STRs reported in this study need further experimental validation, in order to discard any potential artifacts that could exist in both cases and controls, and to confirm their association with PD.

Overall, we have performed the first STR GWAS meta-analysis in PD and reported that STRs contribute to its genetic risk. We have characterized another layer of genetic variation, helping us to gain statistical power to nominate novel candidate PD risk variants and genes, and to provide a more complete reference of the genetic variation that contributes to the disease. Hence this data is a valuable resource for researchers currently dissecting the known PD risk loci. Moving forward, a large-scale GWAS which utilizes calls directly from WGS data and validates hits using long-read sequencing methodologies is essential for fully understanding the contribution of STRs to the genetics of PD.

## METHODS

A summary diagram for the methodological steps followed in the present study is shown in **Supplementary Figure 1**.

### Samples and quality control

All genotyping data was obtained from previously generated IPDGC datasets, consisting of 39,087 individuals (16,642 cases and 22,445 controls) of European ancestry^1^. All individuals provided informed consent for participation in genetics studies, which was approved by the relevant local ethics committee for each of the datasets used. Detailed demographic, sample sizes and PD status are given in **Supplementary Table 1**. Further information along with detailed quality control (QC) methods have been previously published^1,39^. Briefly, for sample QC prior to imputation, individuals with low call rate, discordance between genetic and reported sex, heterozygosity outliers and ancestry outliers were removed. For genotype QC, variants with a missingness rate of > 5%, minor allele frequency (MAF) < 0.01, exhibiting deviations from Hardy–Weinberg Equilibrium (HWE) <1×10^-5^ and palindromic SNPs were excluded.

### STR imputation and filtering

STR genotypes were imputed into the IPDGC SNP unimputed genotyping datasets using Beagle v.5.1^40^ with the 1000 Genomes SNP-STR Haplotype reference panel^17^. In brief, STR genotypes in the reference panel were imputed from STRs called from the catalog-based STR caller hipSTR ^15^ and supplemented using a second STR caller, TREDPARSE^41^. STRs were phased with corresponding SNPs creating a final panel in the 1000 genomes project data that contained 27,185,239 SNP and 445,725 STR markers. Once STRs were imputed into all IPDGC SNP genotype datasets, the STR calls were filtered to facilitate downstream association analysis. First STRs were split from multi-allelic variants to single biallelic variants using the vt variant tool^42^. Finally SNPs and STRs with a dosage R-squared (DR2) <0.3 were removed to filter out low quality imputed variants.

### Study-level STR analysis and meta-analysis

To estimate PD risk, imputed dosages (*i*.*e*. genotype probabilities for a variant to be A/A, A/B, or B/B from 0 to 2) were analyzed using a logistic regression model adjusted for sex, age at onset (AAO) for cases or examination for controls, and the first 10 principal componenets (PCs). To note, AAO could not be included as a covariate for the Myers-Faroud^43^ and Vance (dbGap phs000394) studies as no AAO information was available. Summary statistics were generated using the RVTESTS package^44^ and filtered for a MAF >1%. Meta-analysis was conducted based on the fixed-effect model as implemented in METAL^45^ by combining summary statistics across all 16 IPDGC datasets. All variants with a meta-analysis heterogeneity value of less than 80% (I^2^ <0.80) were kept for further analysis.

### Conditional-joint and linkage disequilibrium analyses

To select candidate variants, we used the Genome-wide Complex Trait Analysis software (GCTA)^19^ to perform conditional and joint analysis (COJO) STRs, from the meta-analysis summary statistics. In order to differentiate associations between STRs and SNPs, we performed two COJO analyses, first with STRs only and second, with STRs and SNPs together. As an LD reference for GCTA we used a sample subset of merged imputed genotypes (hard call threshold of 0.8) from the IPDGC GWAS cohorts^46^, totaling 4,397 PD cases and 9,137 controls. Additionally, we performed LD calculations between top STRs with the previously reported list of 90 PD variants^1^ using PLINK v.1.9^47^ to determine highly linked STRs to known PD risk variants. Hudson plot showing the genome-wide association results for STRs and SNPs separately was done with the *hudson* R package (https://github.com/anastasia-lucas/hudson). Regional plots for the GCTA-nominated independent STRs were done with Locuszoom standalone version^48^.

### eSTR analysis

Using sample level genotypes and gene expression data from the North American Brain Expression Consortium (NABEC)^20^ and the Genotype-Tissue Expression project^21^, we carried out an eQTL analysis with imputed STRs (eSTR). The NABEC data was composed of 343 individuals with genotypes obtained from high-coverage Illumina WGS. Corresponding gene expression data was generated from frontal cortex tissue by RNASeq and normalized gene counts were used. The GTEx v.8 data (dbGaP: phs000424.v7.p2) comprises high-coverage (30X) Illumina WGS data from 838 unrelated samples. We downloaded the fully processed, filtered and normalized gene expression matrices (in BED format) for each of the 13 brain tissues including: amygdala, Anterior cingulate cortex (BA24), caudate (basal ganglia), cerebellar hemisphere, cerebellum, cortex, frontal cortex (BA9), hippocampus, hypothalamus, nucleus accumbens (basal ganglia), putamen (basal ganglia), spinal cord cervical (c-1) and substantia nigra (https://gtexportal.org/home/datasets). WGS genotypes from GTEx and gene start-end coordinates for expression data for GTEx and NABEC were converted from hg38 reference to hg19 using UCSC liftover tool^49^. STRs were imputed as described above. eSTR analysis was performed using the FastQTL software^50^ correcting for PCs 1-10, sample age, sex (if available) and probabilistic estimations of expression residuals factors (PEER) generated using the PEER software^51^: 45 factors for NABEC and 15 for GTEx (as indicated in the GTEx documentation). The 34 top STRs from the meta-analysis along with variants with LD >0.5 (105 STRs total) were used to conduct the eSTR analysis. QQ-plots and box plots were done using ggplot2 R package^52^.

### Heritability estimation

We used the GCTA-LDMS method^19,53^ to estimate the heritability of STRs only, both SNPs and STRs together and SNPs only. The method corrects for LD bias in the estimated variant-based heritability from WGS or imputed data. Heritability estimates and their corresponding standard errors are shown in the liability scale.

### Gene-set, network and pathway enrichment analyses of significant STR loci

To functionally characterize the top associated STRs, we carried out loci connectivity analyses across gene-ontologies and gene-expression datasets using FUMA^24^ and protein-protein interaction networks using Webgestalt^25^. We ran MAGMA gene-wise analysis^23^ using the meta-analysis summary statistics for all STRs, and used the 1000 Genomes SNP-STR dataset as out reference panel^17^. We selected 445 genes with a gene-wise p<0.01 for further analyses (**Supplementary Table 7**). Gene lists were analyzed for functional enrichments using (i) FUMA gene2func tool, (ii) Biogrid PPI Network Topology-based Analysis (NTA) in Webgestalt and (iii) gene property analysis for tissue specificity, using 23,675 genes from Genotype-Tissue Expression (GTEx) RNASeq data^21^ across the 30 general and 54 specific tissues. Data preprocessing and gene expression normalization methods are presented in the FUMA tutorial section (https://fuma.ctglab.nl/tutorial). Bonferroni and Benjamini-Hochberg FDR corrections for multiple testing were performed for MAGMA gene-wise results and functional enrichment analyses, respectively.

## Data Availability

Full STR GWAS summary statistics for the 16 datasets meta-analysed are available at the following link.

https://drive.google.com/file/d/1kD1i6tHdYC5w0xvxWLD4B-bSPqpnwzNV/view?usp=sharing

## Data Availability

Full STR GWAS summary statistics for the 16 datasets meta-analysed are available at https://drive.google.com/file/d/1kD1i6tHdYC5w0xvxWLD4B-bSPqpnwzNV/view?usp=sharing

## Code Availability

The STR imputation, study level GWAS and meta-analysis: https://github.com/neurogenetics/PD_STR_imputation. Downstream analyses: https://github.com/bibb/STR_GWAS_downstream_analysis

## Acknowledgements

We would like to thank all of the subjects who donated their time and biological samples to be a part of this study. We also would like to thank all members of the International Parkinson’s Disease Genomics Consortium (IPDGC). For a complete overview of members, acknowledgements and funding, please see http://pdgenetics.org/partners. This work was supported in part by the Intramural Research Programs of the National Institute of Neurological Disorders and Stroke (NINDS), the National Institute on Aging (NIA), and the National Institute of Environmental Health Sciences both part of the National Institutes of Health, Department of Health and Human Services; project numbers 1ZIA-NS003154, Z01-AG000949-02 and Z01-ES101986. In addition, this work was supported by the Department of Defense (award W81XWH-09-2-0128), and The Michael J Fox Foundation for Parkinson’s Research. This work utilized the computational resources of the NIH HPC Biowulf cluster (http://hpc.nih.gov). The access to part of the participants for this research has been made possible thanks to the Quebec Parkinson’s Network (http://rpq-qpn.ca/en/).

This work was supported by the Simpson Querrey Center for Neurogenetics (to D.K.)

## Conflicts of Interest

D.K. is the Founder and Scientific Advisory Board Chair of Lysosomal Therapeutics Inc. and Vanqua Bio. D.K. serves on the scientific advisory boards of The Silverstein Foundation, Intellia Therapeutics, AcureX and Prevail Therapeutics and is a Venture Partner at OrbiMed. Z.GO. has received consulting fees from Lysosomal Therapeutics Inc., Idorsia, Prevail Therapeutics, Denali, Ono Therapeutics, Neuron23, Handl Therapeutics, Bial Biotech Inc., Deerfield, Lighthouse and Inception Sciences (now Ventus). B.I.B., K.B., C.B., J.R.G., A.B.S. and S.J.L. declare that they have no competing interests.

## Notes

### Clinical Protocols

https://github.com/neurogenetics/PD_STR_imputation.

https://github.com/bibb/STR_GWAS_downstream_analysis

### Author Declarations

Data used in preparation of this article were obtained from controlled access data from the IPDGC, including the Dutch PD GWAS, Finnish PD GWAS, German PD GWAS, Harvard Biomarker Study (HBS), McGill Parkinson's cohort, Myers-Faroud cohort, IPDGC NeuroX cohort, NIA PD GWAS, Oslo PD Study, Parkinson's Disease Biomarker's Program (PDBP), Parkinson's Progression Markers Initiative (PPMI), Baylor College of Medicine / University of Maryland cohort, Spanish Parkinson's part3 cohort, TUBI (Tubingen) cohort, WTCCC PD GWAS and Vance PD cohort. As the analyses in this manuscript utilize secondary analyses of suitably anonymized datasets, they do not require ethics committee review. The respective ethical committees for medical research approved involvement in genetic studies and all participants gave written informed consent in the original publications for the datasets used. The details of these studies can be obtained upon contacting the IPDGC.

