## Supplementary Figure for "Genome-wide contribution of common Short-Tandem Repeats to Parkinson’s Disease genetic risk"

### Supplementary Figures

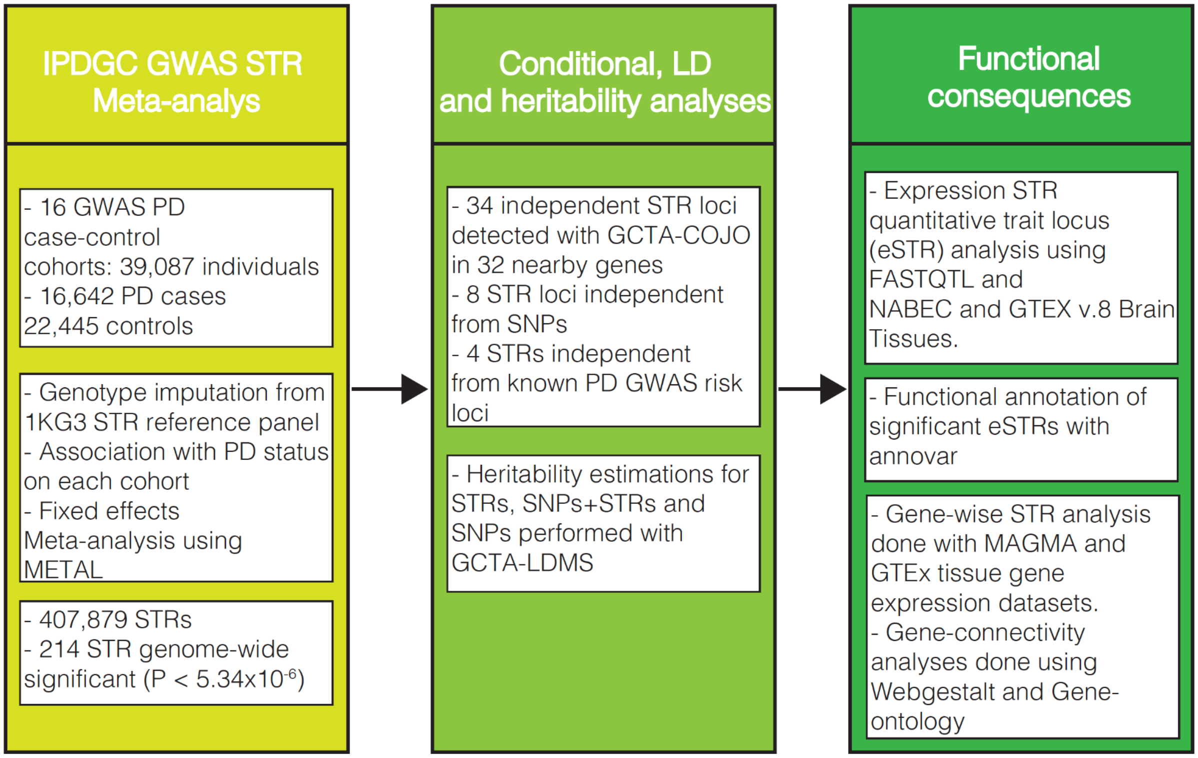


**Supplementary Figure 1. Methodological steps used in the study.** Three main procedures were followed including: i) IPDGC-GWAS cohort imputation with STRs and meta-analysis; ii) Selection of significant and independent STR regions through conditional-joint and LD analyses, and quantifying STR heritability to PD; iii) Functional consequences analyses, including eSTR analysis using gene expression datasets from brain regions, following functional annotation, gene-wise associations and gene connectivity analyses.

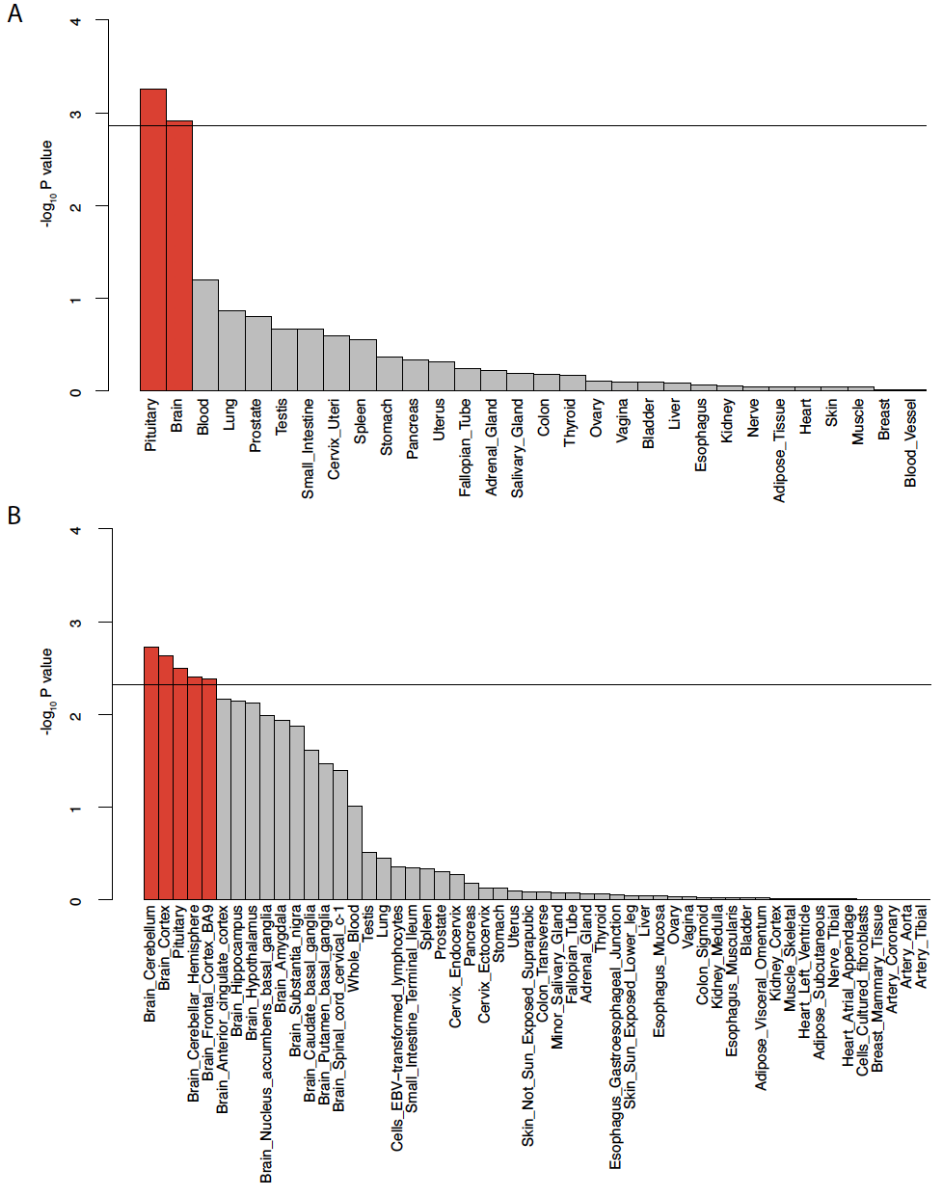


**Supplementary Figure 2. Gene-property enrichment of differentially expressed genes in GTEx general and specific tissues.** MAGMA gene-property enrichment analysis using all genes from the gene-wise enrichment analysis on **(A)** 30 general tissues and **(B)** 54 specific tissues from GTEx v.8. The Y-axis shows the negative logarithm of the enrichment p-value, where bars in red represent significant enrichment surpassing a false discovery rate of 5% (horizontal line).

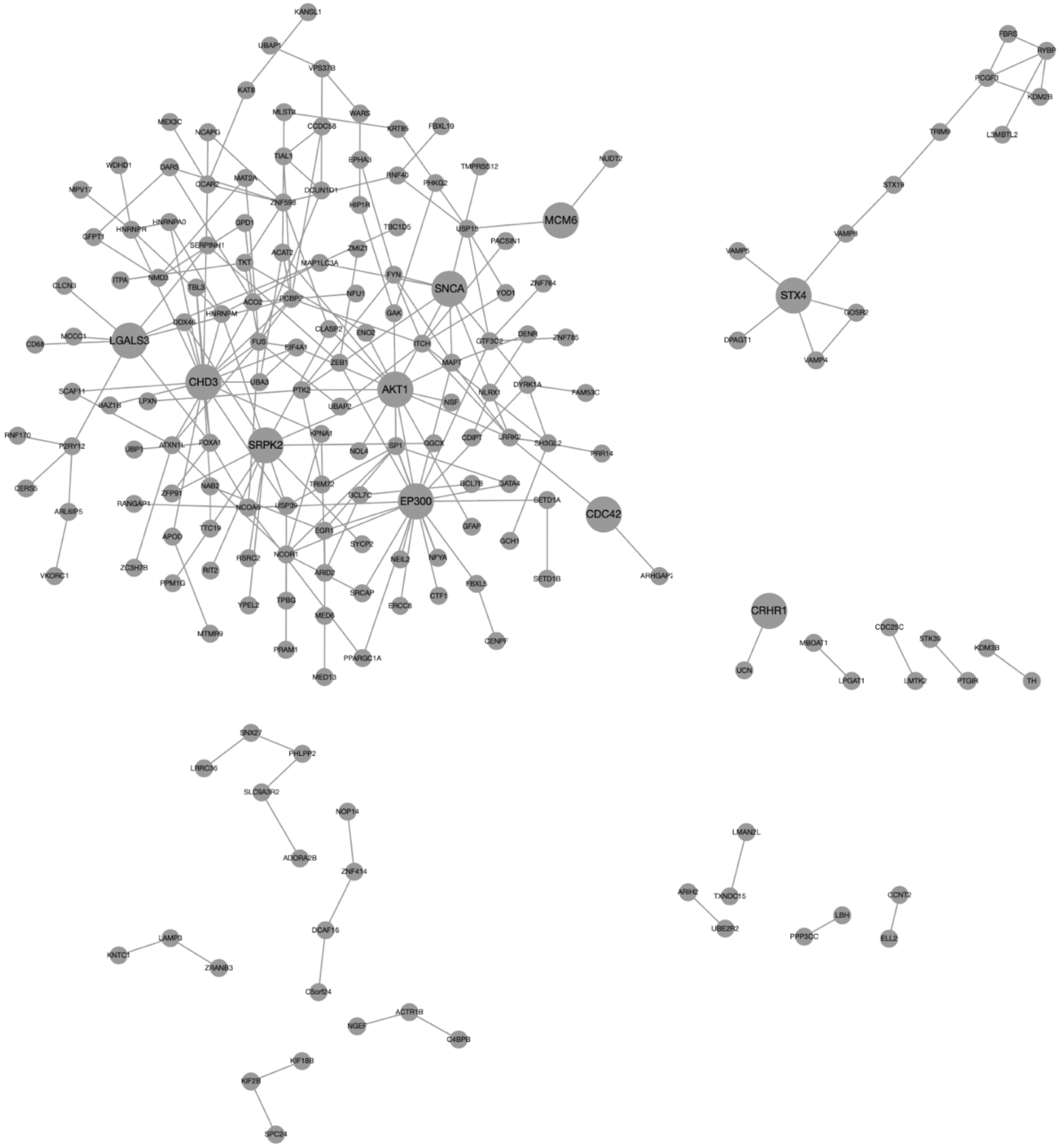


**Supplementary Figure 3. Protein-protein interaction networks obtained from STRs significantly enriched genes.** Sixteen networks obtained from WebGestalt Network Topology Analysis (NTA) using 445 significant genes from MAGMA gene-wise STR enrichment analysis. Each node represents a gene and node size represents top ranked seed genes.
